# Prediction of coronary disease incidence in the general population by circulating RNY(YRNA)-derived small RNA

**DOI:** 10.1101/2020.03.24.20042812

**Authors:** Vera L. Costa, Jean-Bernard Ruidavet, Vanina Bongard, Bertrand Perret, Emanuela Repetto, Maria G. Stathopoulou, Fabrizio Serra, Mohamed Benahmed, Mauduit Claire, Valerie Grandjean, Jean Ferrières, Laurent O. Martinez, Michele Trabucchi

**Affiliations:** Inserm U1065, C3M, Team Control of Gene Expression (10), Université Côte d’Azur, Nice, France; Department of Epidemiology, Health Economics and Public Health, UMR1027 INSERM, Toulouse University, Toulouse University Hospital (CHU), Toulouse, France; Institut National de la Santé et de la Recherche Médicale (INSERM), UMR 1048, Toulouse, France; University of Toulouse, UMR1048, Paul Sabatier University, Toulouse, France; Fédération de Cardiologie, Toulouse University Hospital (CHU), Toulouse, France; Service de Biochimie, Pôle biologie, Hôpital de Purpan, CHU de Toulouse, Toulouse, France

**Keywords:** Y-RNA, atherosclerosis, coronary heart disease, molecular biomarker, primary prevention, prediction

## Abstract

During the development of atherosclerotic lesion, s-RNYs (small RNAs of about 24/34 nucleotides) are derived by the processing of long Ro-associated non-coding RNAs (RNYs) in macrophages. The levels of serum s-RNYs have been found significantly upregulated in patients with coronary heart disease (CHD) compared to age-matched healthy individuals. The present study aimed to examine the predictive value of serum s-RNYs for CHD events in the general population.

Within the frame of nested-case-control study, the GENES study, we measured the absolute expression of a RNY-derived small RNA, the s-RNY1-5p, in the serum of healthy individuals who encountered a CHD event within 12 years of follow-up (n = 31) (Cases) and compared them to individuals who remained event-free (Controls) (n = 30).

The expression of s-RNY1-5p in serum was significantly upregulated in Cases compared to Controls (p = 0.027). The proportion of CHD event-free was significantly higher among individuals with serum s-RNY1-5p below the median value (631 molecule / mL). In a multivariate model adjusted for age, smoking and treatment for hypertension, diabetes and dyslipidemia, the risk of CHD events increased more than 4-fold in individuals with serum s-RNY1-5p above the median value (HR, 4.36; 95%CI, 1.22-15.60). Significant association with CHD events was also observed when considering s-RNY1-5p as a continuous variable (p = 0.022). Serum s-RNY1-5p is an independent predictor of CHD in healthy individuals and could be considered as a biomarker in primary prevention of cardiovascular diseases.

**TRANSLATIONAL PERSPECTIVES:** Here, we reported that s-RNY1-5p was significantly upregulated in the serum of individuals who underwent CHD and was positively associated with CHD events. Those results argue in favor of s-RNY1-5p being a novel predictive molecular biomarker for cardiovascular events. In the future, measurement of s-RNY1-5p expression levels and other 5’ s-RNYs, such as s-RNY4-5p, could be used in clinical practice in addition to classical risk factors to identify those high-risk individuals who might benefit from prevention medicine.

## INTRODUCTION

Atherosclerosis is a progressive disease process that generally begins in childhood and manifests clinically in middle to late adulthood. Biological, clinical, genetic and lifestyle factors represent the major causes of lesion formation, including high level of LDL-C (Low Density Lipoprotein-Cholesterol), low level of HDL-C (High Density Lipoprotein-Cholesterol), high blood pressure, smoking, unhealthy diet, old age, and family history of heart disease. Atherosclerosis starts by the accumulation of atherogenic lipids, including cholesterol from LDL, in the arterial wall, which promotes an inflammatory response by endothelial cells and monocytes/macrophages. In particular, macrophages play a major role in atherosclerosis progression by regulating the inflammatory status of the lesion [1]. In advanced lesions, massive macrophage apoptosis that is not cleared by phagocytic clearance leads to the necrotic plaque formation. The build-up of necrotic plaque ultimately promotes inflammation, lesion disruption, and eventually thrombosis and cardiovascular accident [1]. When this cardiovascular accident occurs in heart arteries is called coronary heart disease (CHD). Therefore, monitoring the degree of macrophage apoptosis in the blood vessels might be a valuable parameter to predict cardiovascular accidents.

Recently, we found that small RNAs of about 24/34 nucleotides (nt) in length derived by the processing of the non-coding YRNAs (RNYs) are significantly upregulated in the serum of patients with CHD compared to age-matched healthy individuals [2]. We have referred to these small RNAs as s-RNYs. RNY genes account for four copies in human genome (RNY1, 3, 4 and 5). Analyses in a cross-sectional study on CHD, comparing 263 CHD-patients with paired controls, positioned s-RNYs as relevant novel independent diagnostic biomarkers for CHD that are associated with atherosclerosis burden [2]. In the context of atherosclerotic lesion and development, serum s-RNYs would be mainly produced by macrophages [2]. RNYs are characterized by extensive base pairing of the 5’ and 3’ regions and by the association with the proteins Ro60 and La/SSB, which are often targeted by the immune system in several autoimmune diseases, including the systemic lupus erythematosus and Sjogren’s syndrome [3]. Circulating s-RNYs are associated with Ro60 protein to form a stable ribonucleoprotein complex that promotes inflammation and apoptosis of monocytes/macrophages by activating the Toll-like receptor (TLR) 7 [4], suggesting that upregulation of serum s-RNYs promotes the atherosclerosis progression.

Here, we tested the hypothesis that measurement of serum s-RNY levels could be useful in the prediction of CHD risk for primary prevention of cardiovascular diseases. To explore this hypothesis, we measured the absolute expression of a sRNYs, the s-RNY1-5p, in the serum of healthy individuals who developed CHD during the 12 years median follow-up from the blood draw and compared them to control individuals who never encountered CHD. Among the different RNY-derived small RNAs previously detected in human serum [2], s-RNY1-5p was selected to study the potential value of RNY-derived small RNAs to predict cardiovascular accidents. In fact, s-RNY1-5p is the most characterized RNY-derived small RNA as marker of CHD status, and like microRNAs, is very stable in the serum [2]. Here, we reported that s-RNY1-5p was significantly upregulated in the serum of individuals who underwent CHD and was positively associated with CHD events. Those results argue in favor of s-RNY1-5p being a novel predictive molecular biomarker for cardiovascular events.

## METHODS

### Sampling Frame

The “Genetique et ENvironnement en Europe du Sud” (GENES) study is a case-control study designed to assess the role of gene-environment interactions in the occurrence of CHD [5]. All participants signed an informed consent form. Biological collection was constituted according to the principles expressed in the Declaration of Helsinki. The study protocol was approved by the local ethics committee (CCPPRB, Toulouse / Sud-Ouest, file #1-99-48, Feb 2000). A collection of biological samples has been constituted (declared as DC-2008-463 #1 to the Ministry of Research and to the regional Health authority) [6]. Control individuals analyzed in the frame of the present work were part of the 834 male individuals from the GENES study, free of CHD and stroke, and aged 45-75 at baseline, randomly selected from the general population using electoral rolls [5]. Those control individuals were followed-up and provided the cases and controls for the nested case-control study of the present work.

### Follow-Up and Case Ascertainment

Control individuals from the GENES study were contacted during the year 2016 either by mail or by telephone and completed self-report questionnaires on clinical events. Among individuals with possible events, clinical information was obtained from medical records in hospital and general practitioners. All available data were gathered concerning hospital admission and type of interventions. Deceased patients’ families and practitioners were contacted to clarify death circumstances, and death certificates were checked to complement clinical and postmortem information. Coronary events (unstable angina, coronary revascularization, non-fatal myocardial infarction), stroke events and the occurrence of peripheral vascular disease were adjudicated by an independent committee. The median follow-up period was 11.9 ± 4.1 years.

### Nested Case-Control Study

In the present work, case patients (n = 31) were individuals who were healthy at baseline and developed either fatal CHD or non-fatal acute coronary events during the follow-up. The control subjects (n = 30), matched for age (± 2 years) and date of recruitment (± 3 months), were participant who remained free of cardiovascular diseases and stroke during follow-up. Analyses were thus based on a nested case-control population of 31 case patients and 30 control participants from the GENES study.

### Baseline Assessment

At the baseline survey, between 2001 and 2004, participants were interviewed and examined in the same health center administered by the national health insurance system. Age, environmental characteristics and information on cardiovascular risk factors were collected through standardized face-to-face interviews, performed by a single physician. Smoking status was classified as current smokers, smokers having quitted for more than 3 years and non-smokers. Alcohol consumption was assessed using a typical week pattern. The total amount of pure alcohol consumption was calculated as the sum of different types of drinks and was expressed as grams per day. Physical activity was investigated through a standardized questionnaire [7] and categorized into two levels, namely, “high” as physical activity for at least 20 min and at least twice a week or “low” physical activity for less than 20 min for once a week or less. Medical history was collected. Presence of dyslipidemia, diabetes mellitus or hypertension was assessed from the subjects’ current treatments. Anthropometric measurements included waist circumference, height and body weight, and body mass index was calculated (BMI, kg/m^2^). Blood pressure and resting heart rate were measured with an automatic sphygmomanometer (OMRON 705 CP). Measurements were performed after a minimum of 5 min rest; average values from two different measurements were recorded for further analysis.

### Laboratory Assays

Blood was collected at inclusion after an overnight fast. Serum samples aliquots were subsequently stored at −80°C until biological analyses. The following biomarkers were assayed with enzymatic reagents on automated analyzers (Hitachi 912 and Cobas 8000, Roche Diagnostics, Meylan, France): serum total cholesterol (TC), HDL-C, triglycerides (TG), fasting glucose, and gamma-glutamyl transferase (γGT). High-sensitive C-reactive protein (hs-CRP) was determined on the same analyzer by immunoturbidimetry assays. Apolipoprotein (Apo) A1, ApoB, and lipoprotein (a) [Lp(a)] were also assayed with an immunoturbidimetric method on an automated analyzer (Roche Diagnostics, France). LDL-C was calculated using the Friedewald formula, with VLDL-cholesterol (VLDL-C) (g/L) = TG (g/L)/5, as long as TG concentration was below 4 g/L [8].

### Absolute quantification of s-RNY1-5p in the serum by quantitative RT-PCR

Absolute quantification of s-RNY1-5p was performed in baseline serum samples from all participants included in the nested case-control study. Before RNA extraction from 0.2 mL human serum, hemolysis was evaluated by measuring through spectrophotometer the oxyhemoglobin at 414 nm and referring to a standard curve made by introducing haemolysis by serially diluting lysed red blood cells in non-haemolysed plasma from a healthy donor. The serum samples showing an optical density (OD) corresponding to > 0.002 % of red blood cells (v/v) were discarded. 0.5 mL of Trizol LS Reagent was added to 0.2 mL of blood serum. Then (*i*) 5 pg of synthetic miRNA-39 from *Caenorhabditis elegans* (miScript miRNA Mimic syn-cel-miR-39-5p, Qiagen) were added as a spike-in control for purification efficiency and (*ii*) 2 μL of glycogen (5 mg/mL) were added to enhance the efficiency of RNA column binding. Purification of extracted total RNA was performed with High Pure miRNA Isolation kit (Roche) according to the manufacturer’s instructions. RNA was eluted in 50 μL of Elution Buffer. The quality of the extracted RNA was checked by ratio between the absorbance values at 260 and 280 nm and between 260 and 230 nm using NanoDrop Technologies ND-1000 spectrophotometer (Supplemental Table 1).

**Table 1.** Baseline characteristics of Controls and Cases when they were first included in the GENES cohort.

|  | Controls (n=30)<br>*Individuals with no coronary event during the follow-up* | Cases (n=31)<br>*Individuals who developed an acute coronary event during the follow-up* | p<sup>a</sup> |
| --- | --- | --- | --- |
| Age (year) | 63.0 (7.8) | 63.8 (8.3) | 0.09 <sup>b</sup> |
| Alcohol, g/day | 18 (18) | 18 (21) | 0.97 |
| Former and current smoker, % | 54.8 | 67.7 | 0.29 <sup>c</sup> |
| Physical activity, high level <sup>d</sup>, % | 29.0 (9) | 32.3 (10) | 0.80 <sup>c</sup> |
| Waist circumference, cm | 96.4 (7.7) | 98.3 (10.4) | 0.44 |
| BMI, kg/m<sup>2</sup> | 26.7 (2.9) | 27.3 (3.6) | 0.48 |
| Fasting glucose, mmol/L | 5.25 (0.89) | 5.59 (0.84) | 0.16 |
| Total cholesterol, g/L | 2.18 (0.31) | 2.29 (0.43) | 0.25 |
| HDL-C, g/L | 0.54 (0.12) | 0.52 (0.10) | 0.47 |
| LDL-C, g/L | 1.43 (0.28) | 1.50 (0.36) | 0.44 |
| Triglycerides, g/L | 1.04 (0.46) | 1.45 (1.02) | 0.03 <sup>e</sup> |
| ApoA-I, g/L | 1.48 (0.22) | 1.49 (0.24) | 0.92 |
| ApoB, g/L | 1.05 (0.19) | 1.12 (0.24) | 0.19 |
| Lp(a), g/L | 0.25 (0.28) | 0.35 (0.39) | 0.32 <sup>e</sup> |
| hs-CRP, mg/L | 4.4 (6.5) | 4.6 (8.6) | 0.38 <sup>e</sup> |
| γGT, IU/L | 45 (41) | 56 (63) | 0.34 <sup>e</sup> |
| Systolic blood pressure, mmHg | 138 (16) | 138 (13) | 0.89 |
| Resting heart rate, beat/min | 62 (9) | 67 (12) | 0.06 |
| Treatment for hypertension, % | 19.4 | 35.5 | 0.14 <sup>c</sup> |
| Treatment for diabetes, % | 9.7 | 6.5 | 0.65 <sup>c</sup> |
| Treatment for dyslipidemia, % | 22.6 | 22.6 | 1 <sup>c</sup> |

*Data are expressed in mean (SD) or %.*
<sup>a</sup> *Paired student's t-test, unless otherwise stated.*
<sup>b</sup> *Wilcoxon signed rank sum test*
<sup>c</sup> *McNemar's test test*
<sup>d</sup> *"high" physical activity during 20 min at least twice a week versus "low" physical activity once a week or less.*
<sup>e</sup> *Analyses performed on log transformed data.*
*BMI: Body Mass Index*
*hs-CRP: high-sensitivity C-Reactive Protein*
*eGFR: estimated Glomerular Filtration Rate*
*γGT: Gamma glutamyl transferase*

Five-fold dilutions of synthetic s-RNY1-5p ranging from 10^9^ to 10^5^ molecules per RT reaction were prepared freshly in Ultrapure water (Invitrogen) before each experiment to create standard curves to use in each quantitative PCR (qPCR) plate. Reverse transcriptase (RT) reaction was performed according to Repetto et al. [2] for the detection of s-RNY1-5p and with TaqMan (Life Technologies) for the cel-miR-39. For s-RNY1-5p detection, RT was performed with the GoScript Reverse Transcriptase Kit (Promega), following the manufacturer’s instructions. Briefly, reagents were freshly mixed before each experiment in a total volume of 20 μL, containing 0.5 mM dNTPs, 0.5 μL reverse transcriptase, 4 μL 5x buffer, 20 units RNase inhibitor, 6.6 μL nuclease-free H2O, 2 μM RT stem-loop primer, and 4 μL of serum RNA or the synthetic s-RNY1-5p. Every step was performed on ice. The reaction was performed at 25 °C for 5 min followed by 42 C for 60 min and 70 °C for 15 min before being held at 4 C. For the synthetic cel-miR-39 detection, RT was performed with the TaqMan microRNA Reverse Transcription Kit (Applied Biosystems), using 1 μL of serum RNA and following the manufacturer’s instructions. The resulting cDNAs were stored at −80 °C, till use for the qPCR experiments.

qPCR amplifications were performed in a total volume of 10 μL, containing 2 μL of freshly prepared RT reaction, 1 μL of a mix containing the specific s-RNY1-5p oligonucleotide primer at 1 μM and the universal oligonucleotide primer at 1 μM (see below for the sequence of the oligonucleotides), 2 μL of nuclease-free H2O, and 5 μl of Fast SYBR Green qPCR Master Mix (Applied Biosystems). Cel-miR-39 detection was performed according to the manufacturer’s instructions (Applied Biosystems) in a final volume of 10 μL. Expression was considered undetectable with threshold cycle (Ct) value ≥ 40. Each sample was run in technical triplicate and a RT negative control without serum RNA was added in each qPCR plate. In addition, in each RT-qPCR plate, we performed s-RNY1-5p standard curve. For each sample, both cel-miR-39 and s-RNY1-5p RT-qPCR reactions were carried out (Supplemental Table 2 and 3).

**Table 2.** sRNY1-5p absolute levels in Controls and Cases when they were first included in the GENES cohort.

|  | Controls (n=30)<br>*Individuals with no coronary event during the follow-up* | Cases (n=31)<br>*Individuals who developed a coronary event during the follow-up* | p |
| --- | --- | --- | --- |
| Mean sRNY1-5p, molecule / mL | 592 (401) | 905 (521) | 0.027 <sup>a</sup> |
| % of sRNY1-5p > median value in the total cohort | 33.3 | 70.0 | 0.022 <sup>b</sup> |

*Data are expressed in mean (SD) or % sRNY1-5p median value in the total cohort = 631 molecules / mL of serum.*
<sup>a</sup> *Paired Student's t-test.*
<sup>b</sup> *McNemar's test.*

**Table 3.** Risk of acute coronary event as a function of sRNY1-5p levels.

|  | sRNY1-5p<br>≤ median <sup>a</sup> | sRNY1-5p<br>> median <sup>a</sup> |  | Per increase of<br>10 molecules / mL |  |
| --- | --- | --- | --- | --- | --- |
|  |  |  | P for<br>trend |  | p |
| HR <sup>b</sup><br>(95% CI) | 1 | 4.36<br>(1.22-15.60) | 0.024 | 1.03<br>(1.01-1.05) | 0.022 |

*HR (95 % CI): Hazard Ratio (95 % confidence interval).*
<sup>a</sup> sRNY1-5p median value = 631 molecules / mL of serum.
<sup>b</sup> Adjusted for age, smoking, treatment for hypertension, diabetes and dyslipidemia.

The cel-miR-39 spike-in measured by RT-qPCR did not vary in all samples (Supplemental Table 2), suggesting an equal yield of RNA purification. These results allowed us to normalize the s-RNY1-5p qPCR data by volume of serum (Supplemental Table 3). Copy number quantification in each sample relied on the standard curve method (Supplemental Figure 1). Standard curve was generated by the StepOne software (ThermoFisher Scientific, MA) based on the concentration of synthetic s-RNY1-5p (Supplemental Figure 1). The automatic threshold function was applied to determine the Ct for each sample and calculate the absolute number of copies. Data were normalized to the initial serum volume (Supplemental Table 3). Primer sequences used in this study are shown in the Supplemental Table 4.

**Figure 1.**
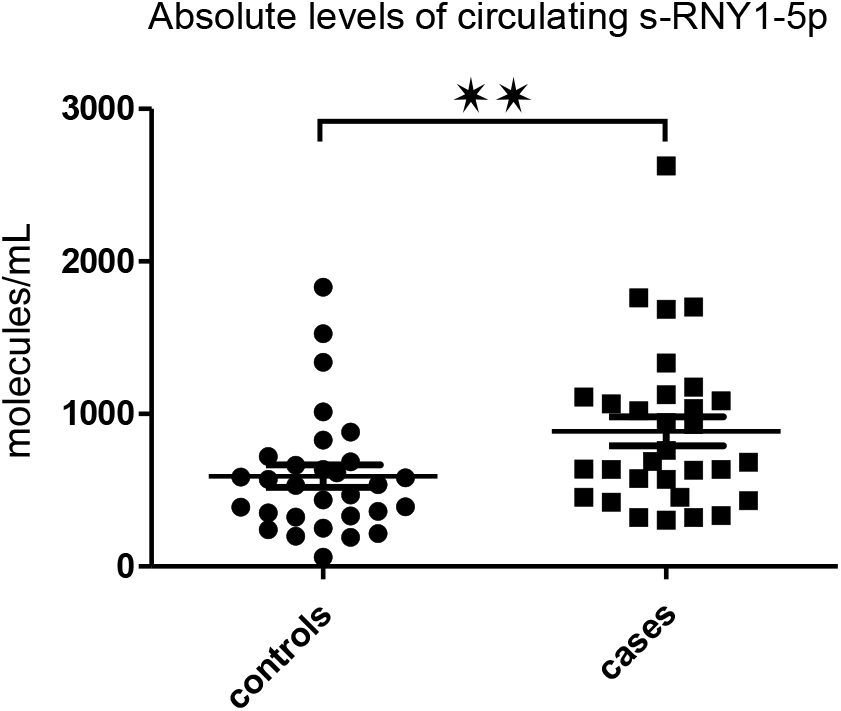
Circulating s-RNY1-5p upregulation in serum of Cases (healthy individuals who developed CHD during a 12-year follow-up period) compared to Controls (individuals who did not developed CHD during a 12-year follow-up period). Box plot showing the absolute expression of circulating s-RNY1-5p in Cases (n =31) and Controls (n = 30). Absolute expression data derived from the standard curves created by synthetic s-RNY1-5p by RT-qPCR, normalized by 1 mL of serum, and presented as median and IQR. Wilcoxon signed rank sum test ** p < 0.01.

### Statistical analysis

Continuous variables are displayed as means and standard deviations (SD) or median and interquartile range (IQR). Categorical variables are presented as proportions. We first described and compared characteristics of participants according to the case-control status. Categorical variables were compared between groups using the McNemar’s test. A logarithmic transformation of variables was performed when necessary. Student’s *t*-test was used to compare the distributions of normally distributed continuous data. Wilcoxon signed rank sum test was used when distribution departed from normality, or when homoscedasticity was rejected.

Cumulative risk of patients for coronary event (fatal and non-fatal) was determined by the Kaplan-Meier method and compared using the Log-rank test for the individual endpoints of coronary events.

The relation between baseline levels of s-RNY1-5p and the case-control status was assessed using Cox proportional hazards regression analysis. Cox regression analyses were performed with adjustment on classical cardiovascular risk factors (age, smoking, treatments for dyslipidemia, hypertension and diabetes). We tested the proportionality assumption using cumulative sums of martingale-based residuals. We performed regression analysis with polynomial models (quadratic and cubic) to examine for possible non-linear relationship between s-RNY1-5p and occurrence of coronary event.

All statistical analyses were carried out using the SAS statistical software package 9.4 (SAS Institute, Cary, NC) or STATA statistical software, release 14.2 (STATA Corp, College Station, TX). All tests were two-sided and were considered significant at a p value < 0.05.

## RESULTS

The Nested Case-Control study we used here, comprised 61 men from the GENES study, apparently healthy at baseline. The median follow-up period was 11.9 ± 4.1 years. Case patients (n = 31) were individuals who develop fatal (n = 16) or non-fatal acute coronary syndrome such as unstable angina and non-fatal myocardial infarction (n = 15) during the follow-up. The control subjects (n = 30), matched for age (± 2 years) and date of recruitment (± 3 months), were participants who remained free of cardiovascular diseases and stroke during follow-up. Comparison of individual’s data at the recruitment in the cohort is given in Table 1. Results are presented distinguishing two groups: the ‘‘Controls group’’ (individuals with no coronary event during the follow-up) and the ‘‘Cases group’’ (individuals who developed an acute coronary event during the follow-up). There was no significant differences between biochemical and physiological variables among the Cases and Controls, except for TG levels that were significantly higher in Cases (p = 0.03). No significant difference between Cases and Controls was noted with regards to treatment for dyslipidemia and diabetes. The proportion of Cases treated for hypertension tended to be higher than in Controls (19.4 % versus 32.3 %, p = 0.14). Overall, the present cohort population was representative of the general male population of this age range in the early 2000s in Southern France in term of biological variables and treatments [9].

Absolute values of serum s-RNY1-5p expression at baseline was measured by RT-qPCR assay using total RNA extracted from the Nested Case-Control study. As shown in Figure 1 and Table 2, expression levels of serum s-RNY1-5p, expressed in number of molecules per mL, were significantly higher in Cases. Accordingly, the percentage of individuals with s-RNY1-5p above the median value (631 molecules / mL) was also significantly higher in Cases compared to Controls (70.0% *versus* 33.3%, p = 0.022). The relation between baseline levels of s-RNY1-5p and the case-control status was assessed using Cox proportional hazards regression analysis. As reported in Table 3, in a multivariate model adjusted for classical risk factors (age, smoking and treatment for hypertension, diabetes and dyslipidemia), the risk of CHD event increased more than 4-fold in individuals with serum s-RNY1-5p above the median value (Hazard Ratio (HR), 4.36; 95%CI, 1.22-15.60). Significant association with CHD events was also observed when considering s-RNY1-5p as a continuous variable. Indeed, the adjusted HR for coronary event per 1-SD increase in s-RNY1-5p was 1.03 (95% CI, 1.01-1.05, p = 0.022).

Then, cumulative risk of patients for coronary event (fatal and non-fatal) was determined by the Kaplan-Meier method according to the median s-RNY1-5p value (631 molecules / mL) and compared, using the Log-rank test for the individual endpoints of CHD events (Figure 2). This analysis indicated that the proportion of CHD event-free individuals was significantly lower in individuals with s-RNY1-5p level above the median value (Figure 2, Log-rank test: CHI2=4.8, p = 0.028). Thus, high levels of s-RNY1-5p are associated with an increased risk of developing a first coronary event.

**Figure 2.**
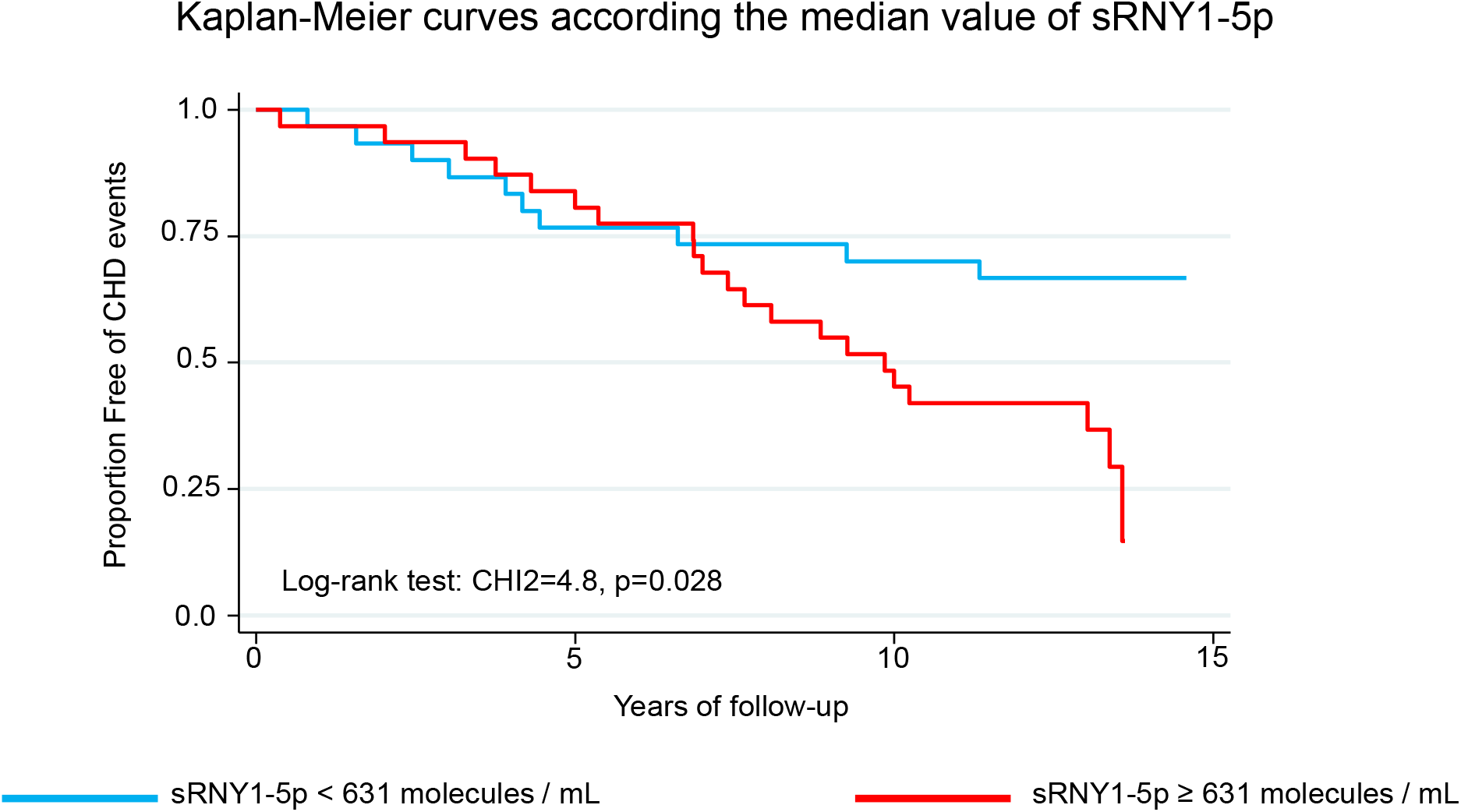
Predictive value of serum s-RNY1-5p for CHD events. Kaplan-Meier probability curves to encounter CHD events according to s-RNY1-5p level, above or below the median value (631 molecule /mL).

## DISCUSSION

s-RNYs-5p represent a large proportion of all small RNAs expressed in human serum and plasma [10]. They circulate as part of a complex with a mass between 100 and 300 kDa [10], which contains the protein Ro60 [2]. Specific subtypes of s-RNYs-5p have been found dysregulated in the serum of patients with breast cancer and have been proposed as novel biomarkers for diagnosis in this pathology [11], as well in head and neck squamous cell carcinoma [12]. In 2015, we found that s-RNY1-5p is produced by macrophages and that its relative expression levels in serum, normalized by endogenous or spike-in controls with the 2-ΔΔCt method, was significantly upregulated in CHD patients compared to age-matched healthy individuals [2]. The goal of the present study was to evaluate whether serum s-RNY levels could be useful for primary prevention of CHD. To this aim, for the first time, we have developed and validated a method to quantify the absolute expression of s-RNY1-5p in human serum. Based on a Nested Case-Control Study of apparently healthy individuals at baseline and followed for a median duration of 12 years, we observed that serum s-RNY1-5p levels are higher in individuals who developed an acute coronary event during the follow-up. Interestingly, we observed that s-RNY1-5p levels were positively associated to CHD events, independently of classical risk factors.

Among the small non-coding RNAs, microRNAs (miRNAs) have also become potential biomarkers for human pathologies, including cardiovascular disorders [13]. miRNAs are ∼20 nucleotides long endogenous non-coding RNAs abundantly expressed in virtually all human cells and associated with AGO proteins to repress gene expression at the post-transcriptional level [14]. miRNAs are localized in mostly all cellular organelles and are also present in the extracellular environment [15]. More than 100 miRNAs have been detected in serum and designed as circulating miRNAs [13]. Because of their extracellular stability, measurement of circulating miRNA dysregulation has been proposed as novel method for the diagnosis of several human pathologies, including CHD [13]. Among them, elevated levels of miR-499 seem to be associated with unstable angina and myocardial infarction within 3 hours of symptom onset, supporting a role of miR-499 as potential biomarker for early diagnosis of CHD [16]. However, to date, none of the dysregulated circulating miRNAs may predict cardiovascular accidents in long-range forecast. To the best of our knowledge, the present study is the first one demonstrating the potential role of a small non-coding RNA in predicting CHD development, underlining therefore the originality and importance of our findings.

The prevalence of CHD is increasing and the burden on the healthcare system stems from late diagnosis. In this context, the importance of early identification and preventative intervention is being recognized worldwide. The recommended method for risk screening according to the European Society of Cardiology (ESC) Guidelines for the diagnosis and management of chronic coronary syndromes is the Systematic COronary Risk Evaluation (SCORE) [17]. SCORE assesses a combination of clinical parameters (age, gender, smoking status, systolic blood pressure) and blood-based biomarkers (lipid panel of total cholesterol, HDL-C, LDL-C and TG) to estimate a 10-year risk of fatal CHD. However, both the ECS and American College of Cardiology/American Heart Association (ACC/AHA) recognize that current risk evaluation methods are unreliable and have limitations. The strongest pain point experienced by the healthcare system is diagnosis of patients classified as ‘moderate risk’ following SCORE evaluation. These patients represent a large portion of the population, commonly those who are 35 to 50 years and diabetic. Primary care physicians then need to reclassify these patients as high or low risk by referring them to clinics or hospitals for expensive cardiac diagnostic testing. As a result, often the prevalence of CHD is overestimated, wasting costly and strained healthcare resources. Therefore, a more accurate biomarker that complements the lipid panel and SCORE assessment would improve the overall predictive performance of CHD risk evaluation. In this respect, our present results indicated that sRNYs, and particularly s-RNY1-5p, could address this unmet need for an early non-invasive biomarker that increase accuracy of CHD risk assessment.

One of the strengths of our study was the use of a matched control group of individuals who did not develop CHD during the follow-up period. However, several limitations of the present study must be noted. Firstly, the small sample size of the study is a limitation and further measurements are foreseen within the framework of a large prospective cohort to establish more firmly the predictive value of s-RNY1-5p determinations. Yet, our study is the first to report the positive association between s-RNY1-5p upregulated levels with the prediction of CHD events. Secondly, our study was designed only with men, which has the advantage of recording a larger number of events than in a mixed all-gender cohort. Particularly, this cohort is composed of participants living in Toulouse area (South-west France), in which there is low cardiovascular mortality rate in women before 65 years of age [18], thus limiting the translatability of our results to women.

Despite these limitations, the present study represents a step forward towards a critical assessment of circulating small non-coding RNAs as biomarkers for prediction of CHD. In the future, measurement of s-RNY1-5p expression levels and other 5’ s-RNY, such as s-RNY4-5p, could be used in clinical practice in addition to classical risk factors to identify those high-risk individuals who might benefit from prevention medicine.

## Data Availability

all raw data in the manuscript are present in this submission

## ACKNOWLEDGEMENT

We are indebted to C. Lorente for technical help.

## SUPPLEMENTAL INFORMATION

## Supplemental Figure legends

**Supplemental Figure 1. s-RNY1-5p standard curve generated by using synthetic RNA oligonucleotides and RT-qPCR**. Illustrative s-RNY1-5p standard curve obtained in one qPCR plaque run together with 24 serum samples (in blue) represent the amount obtained for 16 μL.

## Supplemental Table legends

**Supplemental Table 1: Quantification of serum RNA**

**Supplemental Table 2: Quantification of cel-miR39 by RT-qPCR**

**Supplemental Table 3: Quantification of s-RNY1-5p molecules per mL by RT-qPCR**

**Supplemental Table 4: Primer sequences**

